# Differential Risk of SARS-CoV-2 Infection by Occupation: Evidence from the Virus Watch prospective cohort study in England and Wales

**DOI:** 10.1101/2021.12.14.21267460

**Authors:** Sarah Beale, Susan Hoskins, Thomas Byrne, Wing Lam Erica Fong, Ellen Fragaszy, Cyril Geismar, Jana Kovar, Annalan M D Navaratnam, Vincent Nguyen, Parth Patel, Alexei Yavlinsky, Anne M Johnson, Martie Van Tongeren, Robert W Aldridge, Andrew Hayward, the Virus Watch Collaborative

## Abstract

**Background:** Workers differ in their risk of SARS-CoV-2 infection according to their occupation, but the direct contribution of occupation to this relationship is unclear. This study aimed to investigate how infection risk differed across occupational groups in England and Wales up to April 2022, after adjustment for potential confounding and stratification by pandemic phase.

**Methods:** Data from 15,190 employed/self-employed participants in the Virus Watch prospective cohort study were used to generate risk ratios for virologically- or serologically-confirmed SARS-CoV-2 infection using robust Poisson regression, adjusting for socio-demographic and health-related factors and non-work public activities. We calculated attributable fractions (AF) amongst the exposed for belonging to each occupational group based on adjusted risk ratios (aRR).

**Findings:** Increased risk was seen in nurses (aRR=1.44, 1.25-1.65; AF=30%, 20-39%), doctors (aRR=1.33, 1.08-1.65; AF=25%, 7-39%), carers (1.45, 1.19-1.76; AF=31%, 16-43%), primary school teachers (aRR=1.67, 1.42-1.96; AF=40%, 30-49%), secondary school teachers (aRR=1.48, 1.26-1.72; AF=32%, 21-42%), and teaching support occupations (aRR=1.42, 1.23-1.64; AF=29%, 18-39%) compared to office-based professional occupations. Differential risk was apparent in the earlier phases (Feb 2020 - May 2021) and attenuated later (June - October 2021) for most groups, although teachers and teaching support workers demonstrated persistently elevated risk across waves.

**Interpretation:** Occupational differentials in SARS-CoV-2 infection risk vary over time and are robust to adjustment for socio-demographic, health-related, and non-workplace activity-related potential confounders. Direct investigation into workplace factors underlying elevated risk and how these change over time is needed to inform occupational health interventions.

## Introduction

Notable occupational inequalities in infection risk have emerged during the Coronavirus Disease 2019 (COVID-19) pandemic. Research and surveillance data across various global regions have repeatedly indicated elevated risk of severe acute respiratory syndrome coronavirus 2 (SARS-CoV-2) infection in workers in various essential and/or public-facing industries, such as health and social care, transportation, education, and cleaning and service occupations ^1 2 3 4 5^ compared to other workers or the adult population. Occupational differences in the ability to work from home, the frequency and intensity of workplace exposure to other people, environmental features of the workspace, and the implementation of infection control procedures plausibly contribute to differential risk of infection and transmission at work ^6 7 8^. However, occupation is intimately linked with other socio-demographic factors such as deprivation, household size, activities outside the workplace and health status, that can compound to influence infection risk^9 10^. Establishing the contribution of work-related exposure to occupational inequalities in infection risk consequently depends on careful consideration of other non-occupational factors.

Few estimates of the effect of occupation on SARS-CoV-2 infection risk or outcomes have comprehensively accounted for sociodemographic confounding beyond age and sex. Age, sex, geographic factors, education, living conditions, and pre-pandemic health were estimated to account for 70-80% of the effect of occupation on COVID-19 mortality in the UK in 2020^11^. Healthcare, care, and some service and transport occupations (among men) and elementary cleaning and plant workers (among women) demonstrated elevated mortality compared to all other occupations, but the strength of these estimates was greatly attenuated by adjustment. While these findings indicate the importance of comprehensive adjustment, mortality data are strongly affected by clinical risk factors and the impact of work-related factors on differential infection risk cannot therefore be inferred from these findings.

Data from Germany^12^ (February – September 2020) and Sweden^13^ (January 2020 – February 2021) indicates elevated risk of infection amongst essential workers – including health, care, and service workers – compared to non-essential workers across the respective study periods, after adjustment for a range of socio-demographic factors. However, occupational differences in risk may vary by global region and comparative investigation for the UK is limited. Probability of antigen test positivity differed little across occupations after adjustment for age, sex, region, ethnicity, household composition, deprivation, ability to work from home, use of face coverings at work, and ability to socially distance at work, based on the UK Office for National Statistics (ONS) Coronavirus Infection Survey^14^ between early September-early January 2021. However, the inclusion of work-related potential mediators in this analysis precludes disaggregating the impact of occupational and non-occupational factors.

Differential risk across occupations is also plausibly influenced by time, due to changes in public health interventions and restrictions - including sectoral closures, social distancing, and infection control in the workplace - as well as fluctuating levels of community transmission across the pandemic and changes in immunity due to infection or vaccination. Preliminary evidence from the UK and Norway suggests that occupational differences in infection risk vary across time, with health ^3,15,16,17^ and social care workers ^15,16^ and transport workers ^3^ demonstrating elevated infection risk during the first pandemic wave and other public-facing occupations including education ^15,16^, manufacturing ^15,16^ and food service as well as transport workers ^3^ demonstrating elevated risk in the second wave. More recent data including the period of relaxation of pandemic restrictions in the UK are lacking, as are estimates over time comprehensively adjusted for non-occupational factors.

Using data from a prospective community cohort study in England and Wales (Virus Watch) ^18^, this study aimed to extend current understanding of the direct effect of occupation on SARS-CoV-2 infection risk over time. Specific objectives were: (1) to estimate the relative risk of SARS-CoV-2 infection by occupation across the pandemic, adjusting for socio-demographic and health-related factors and non-work public activities; (2) to investigate whether occupational infection risk differed across pandemic waves; and (3) to estimate the attributable fraction amongst the exposed for different occupations overall and by pandemic wave.

## Methods

### Ethics Approval

Virus Watch was approved by the Hampstead NHS Health Research Authority Ethics Committee: 20/HRA/2320, and conformed to the ethical standards set out in the Declaration of Helsinki. All participants provided informed consent for all aspects of the study.

### Participants

Participants in the current study (*n*=15,190) were an adult sub-cohort of the Virus Watch longitudinal cohort study (*n*=58,692 as of 12/02/2022 when cohort recruitment was completed). Participants were included in the present study if they were (1) ≥16 years, (2) in employment or self-employment and reported their occupation upon study registration, and (3) completed at least one monthly survey between November 2020 and March 2022 concerning their activities across a recent week. Further detail of the full Virus Watch cohort study, including inclusion criteria for the full cohort, can be obtained from the study protocol^18^.

### Exposure

Occupation was derived based on free-text responses to the Virus Watch baseline survey (94% of classified responses) or a Virus Watch monthly survey conducted in February 2022 (6% of classified responses); the baseline survey was used as a preferential source, with the monthly survey used only if participants’ occupation was missing at baseline. Following the protocol recommended by the UK Office for National Statistics (ONS)^19^, we performed semi-automatic coding using Cascot Version 5.6.320 to assign participants UK Standard Occupational Classification (SOC) 2020 codes^19^. Occupations were then classified into the following groups, which aimed to broadly reflect workplace environments while retaining, as far as possible, ONS-defined occupational skill groupings: administrative and secretarial occupations; healthcare occupations; indoor trade, process & plant occupations; leisure and personal service occupations; managers, directors, and senior officials; outdoor trade occupations; sales and customer service occupations; social care and community protective services; teaching education and childcare occupations; transport and mobile machine operatives; and other professional and associate occupations (broadly office-based professional and associate professional occupations).

Where possible, we also extracted more specific occupational groupings based on three-digit SOC groups for occupations within the essential worker classification^21^ and classified by the investigators as public facing/frontline roles. These more detailed occupational groups were included where group sizes exceeded *n*=100 and some SOC groupings were split or combined together to reflect working environment/role, to yield the following included groupings: nurses, doctors, warehouse and process/plant occupations, food preparation and hospitality occupations, teachers (primary school), teachers (secondary school), teachers (higher education), teaching assistants and support occupations, carers, social work and welfare occupations, cleaners, and salespeople/cashiers/shopkeepers.

For further methodological details of exposure classification and UK SOC 2020 codes within each category, please see ‘Occupational Classification’ in the Supplementary Materials.

### Outcomes

The outcome of interest was binary SARS-CoV-2 infection status (yes/no ever infected) based on any clinical evidence of infection (positive lateral flow (LFT), polymerase chain reaction (PCR), anti-nucleocapsid antibody serological test, or anti-spike antibody serological test in absence of vaccination). Participants were censored after first infection, as susceptibility to reinfections was not the focus of this paper. Please see ‘Clinical Outcomes’ in the Supplementary Material for further information about clinical data in Virus Watch and how infection status was derived.

Where possible, we attributed results to either the earlier phase of the pandemic characterised by stringent public health restrictions and the dominance of the SARS-CoV-2 wild type and subsequently Alpha variant in the UK (comprising Wave 1 and 2 between February 2020 to May 2021), the mid phase characterised by relaxation of restrictions and the dominance of the Delta variant (comprising Wave 3 from June 2021 to November 2021), or the later phase characterised by further relaxations of restrictions and the dominance of the Omicron variant (comprising Wave 4 from December 2021 to April 2022) based on test date. Test results were only available until 1 April 2022 due to the termination of the national testing programme in England affecting self-reported testing data and the termination of monthly serological testing in Virus Watch. Waves 1 and 2 were amalgamated into a single phase as it was not possible to attribute specific waves to serology tests conducted during Wave 2, and as mass population testing was largely introduced after the first pandemic wave in England and Wales. Both Waves 1 and 2 included periods of stringent public health restrictions, whilst Waves 3 and 4 occurred during the relaxation of public health measures in included regions, with a brief reintroduction of some limited restrictions in December 2021 and January 2022 due to the emergence of the Omicron variant. Some infections could not be attributed to a particular period as they were based on seropositivity without a prior seronegative result.

### Covariates

Where appropriate (see Statistical Analyses), models were adjusted for the following socio-demographic and health-related covariates: age (<30, 30-39, 40-49, 50-59, 60+ years), sex at birth, binary vulnerability status (defined as any condition on the UK NHS/government list of clinically vulnerable conditions ^22^, obesity, and/or having received an NHS shielding letter), minority ethnicity (White British vs other), geographic region (ONS national region), deprivation based on English or Welsh Indices of Multiple Deprivation Quintile derived from postcode, annual household income (£0-24,900, £25,000-£49,999, £50,000-£75,000, and £75,000+) and household size (excluding participant).

Models were adjusted for non-work public activities based on monthly surveys where participants reported the median number of days that they undertook the following activities across each survey week: using transport (using a bus, underground or overground train/tram, taxi, or sharing a car with a non-household member), visiting essential shops, and leisure and social activities (attending the theatre, cinema, concert or sports event; eating in a restaurant, cafe or canteen; going to a bar, pub or club; going to a party; or non-essential shops or personal care services). Responses from November 2020 and February - April 2021 were allocated to Waves 1 and 2, with the second wave used to extrapolate to both early phases of the pandemic. Responses from May 2021-October 2021 were allocated to Wave 3, and from November 2021 – March 2022 to Wave 4. Monthly surveys were conducted towards the end of each month, so surveys conducted on the boundary months between pandemic waves were allocated to the subsequent wave.

### Statistical Analyses

To assess the influence of occupation on SARS-CoV-2 infection risk, we performed Poisson regression with robust standard errors, an established method to estimate risk ratios for binary outcomes ^23^. Separate models were conducted for the full pandemic and by wave, with the reference category set as (1) ‘Other Professional and Associate Occupations’, the largest occupational group in Virus Watch broadly comprising office-based professional occupations (see Supplementary Table S1) with a low absolute infection risk (see Supplementary Table S2), and (2) the full working population of Virus Watch excluding the occupational group under consideration. We identified potential confounders based on a purpose-developed directed acyclic graph (DAG - see ‘Directed Acyclic Graphs’ in Supplementary Materials), with models presented unadjusted and fully adjusted for the following confounders according to our DAG: age, sex, ethnicity, region, deprivation and household size, vulnerability status, and non-work public activities. Vaccination status was not directly included in models due to the inclusion of variables determining vaccination (i.e., age, health status, and occupation in the case of vaccination) due to UK protocols and related position on the causal pathway between occupation and infection risk (see DAGs in Supplementary Figures 1a and 1b). No evidence of multicollinearity emerged based on variance inflation factors for any model. We performed a sensitivity analysis limited to participants who had undergone serological testing (*n*=9114) to address potential differential access and testing behaviour for virological/antigen testing across occupations; it was only possible to perform this analysis on broad occupational groups across the full study period, and not for specific occupations or by wave due to limited statistical power (see ‘Clinical Outcomes’ in the Supplementary Materials).

Based on the fully-adjusted models, we calculated attributable fractions for the exposed subpopulations (AFs) using the *punaf* programme in Stata Version 16 ^24^. Attributable fractions range from -∞ to 1, with negative values indicating a protective effect and positive values indicating a harmful effect ^24^; while negative values are often transformed to express cases prevented in the unexposed group, we did not transform estimates in order to facilitate comparison by leaving all estimates with the same denominator.

Missing data were limited for all included sociodemographic variables (0-6%) and complete cases were included in the final analyses. We conducted a missing data sensitivity analysis by applying multivariate imputation by chained equations (*mice* package in R Version 4.0.3^25^) with 5 datasets with 50 iterations per dataset to socio-demographic variables and re-testing models.

## Results

Selection of participants into the current study based on inclusion criteria presented in Figure 1, with demographic features of included participants (*n*=15,190) reported in Table 1.

**Table 1.**
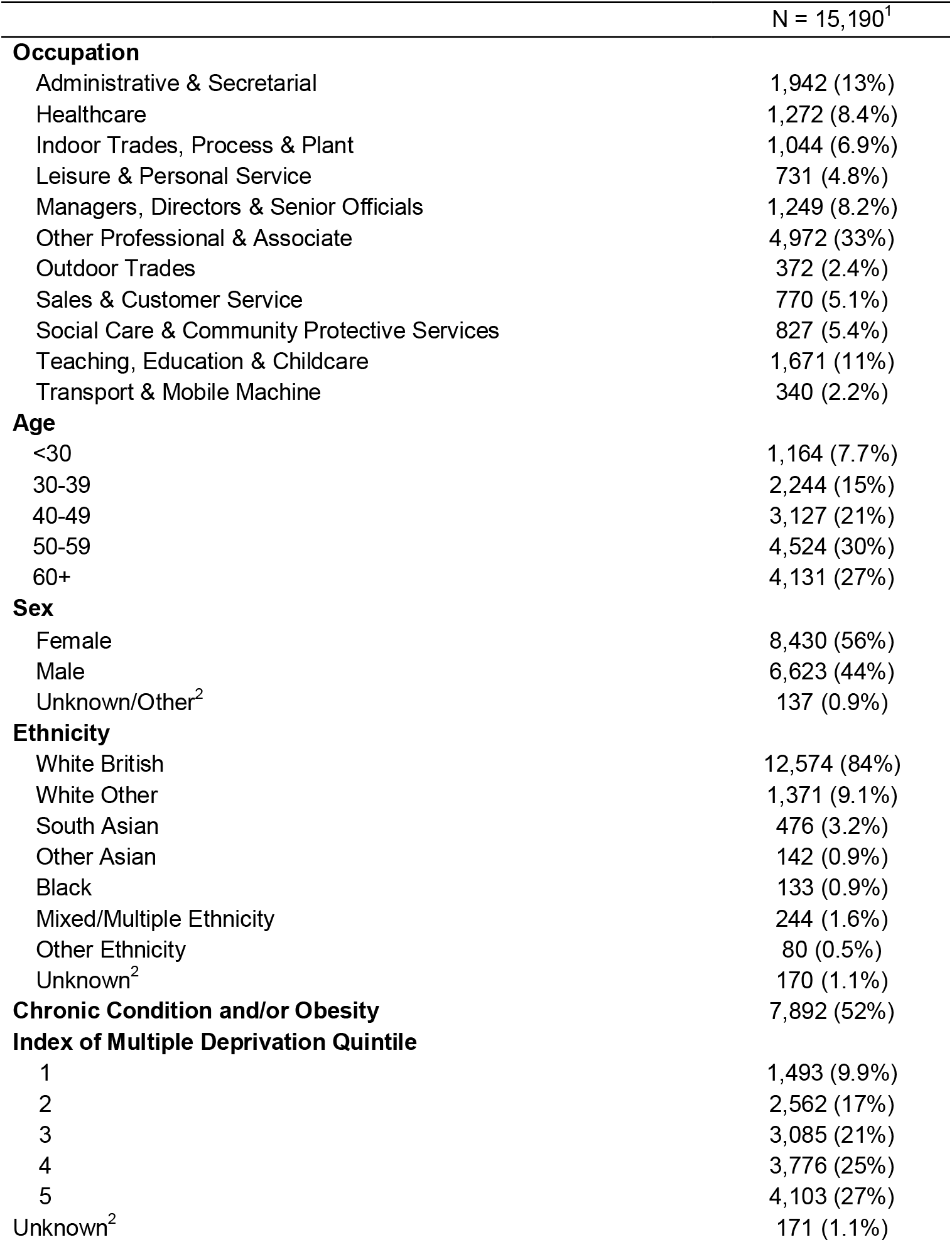
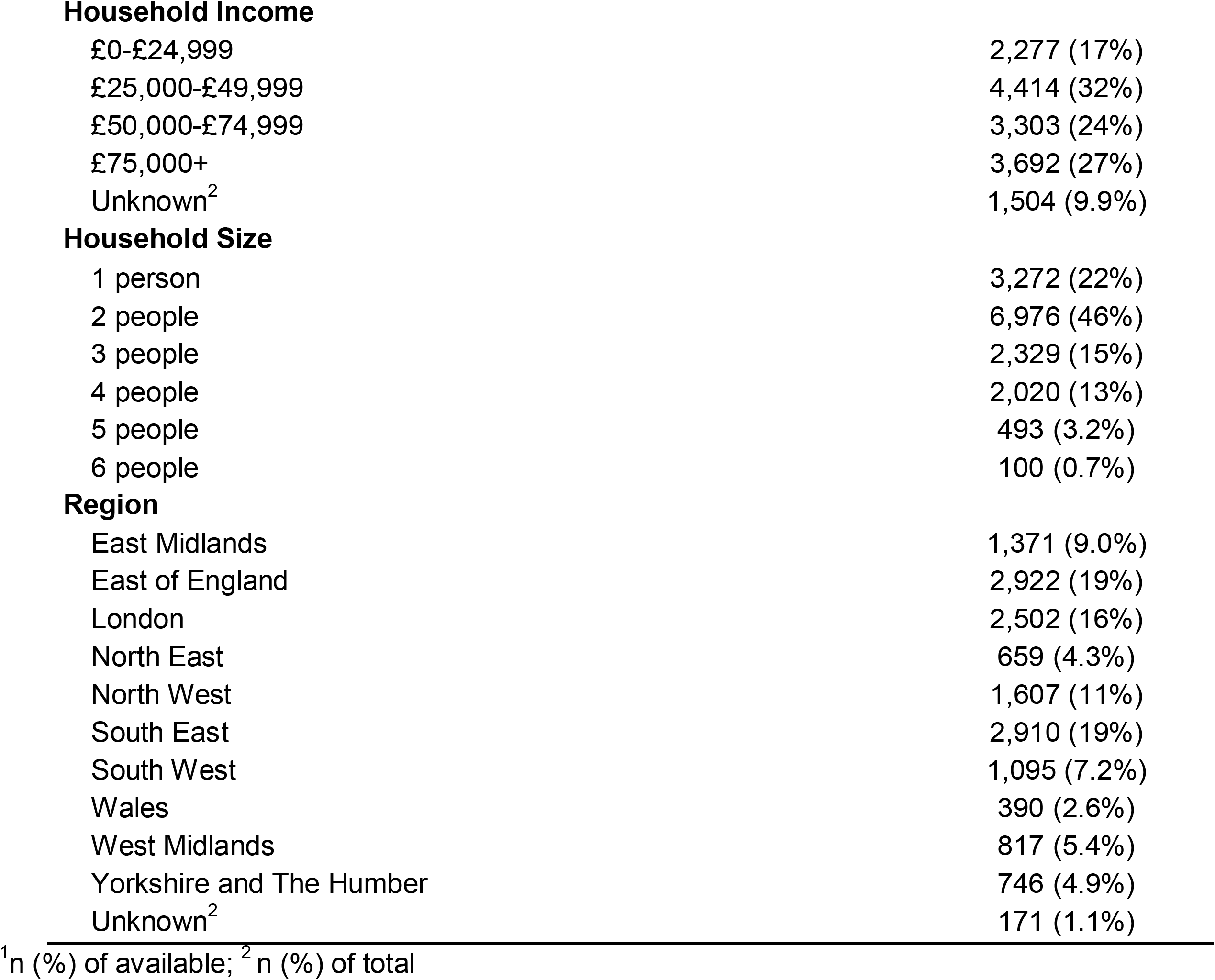
Characteristics of Study Participants

|  | N = 15,190<sup>1</sup> |
| --- | --- |
| **Occupation** |  |
| Administrative & Secretarial | 1,942 (13%) |
| Healthcare | 1,272 (8.4%) |
| Indoor Trades, Process & Plant | 1,044 (6.9%) |
| Leisure & Personal Service | 731 (4.8%) |
| Managers, Directors & Senior Officials | 1,249 (8.2%) |
| Other Professional & Associate | 4,972 (33%) |
| Outdoor Trades | 372 (2.4%) |
| Sales & Customer Service | 770 (5.1%) |
| Social Care & Community Protective Services | 827 (5.4%) |
| Teaching, Education & Childcare | 1,671 (11%) |
| Transport & Mobile Machine | 340 (2.2%) |
| **Age** |  |
| <30 | 1,164 (7.7%) |
| 30-39 | 2,244 (15%) |
| 40-49 | 3,127 (21%) |
| 50-59 | 4,524 (30%) |
| 60+ | 4,131 (27%) |
| **Sex** |  |
| Female | 8,430 (56%) |
| Male | 6,623 (44%) |
| Unknown/Other<sup>2</sup> | 137 (0.9%) |
| **Ethnicity** |  |
| White British | 12,574 (84%) |
| White Other | 1,371 (9.1%) |
| South Asian | 476 (3.2%) |
| Other Asian | 142 (0.9%) |
| Black | 133 (0.9%) |
| Mixed/Multiple Ethnicity | 244 (1.6%) |
| Other Ethnicity | 80 (0.5%) |
| Unknown<sup>2</sup> | 170 (1.1%) |
| **Chronic Condition and/or Obesity** | 7,892 (52%) |
| **Index of Multiple Deprivation Quintile** |  |
| 1 | 1,493 (9.9%) |
| 2 | 2,562 (17%) |
| 3 | 3,085 (21%) |
| 4 | 3,776 (25%) |
| 5 | 4,103 (27%) |
| Unknown<sup>2</sup> | 171 (1.1%) |
| **Household Income** |  |
| £0-£24,999 | 2,277 (17%) |
| £25,000-£49,999 | 4,414 (32%) |
| £50,000-£74,999 | 3,303 (24%) |
| £75,000+ | 3,692 (27%) |
| Unknown<sup>2</sup> | 1,504 (9.9%) |
| **Household Size** |  |
| 1 person | 3,272 (22%) |
| 2 people | 6,976 (46%) |
| 3 people | 2,329 (15%) |
| 4 people | 2,020 (13%) |
| 5 people | 493 (3.2%) |
| 6 people | 100 (0.7%) |
| **Region** |  |
| East Midlands | 1,371 (9.0%) |
| East of England | 2,922 (19%) |
| London | 2,502 (16%) |
| North East | 659 (4.3%) |
| North West | 1,607 (11%) |
| South East | 2,910 (19%) |
| South West | 1,095 (7.2%) |
| Wales | 390 (2.6%) |
| West Midlands | 817 (5.4%) |
| Yorkshire and The Humber | 746 (4.9%) |
| Unknown<sup>2</sup> | 171 (1.1%) |

<sup>1</sup>n (%) of available; <sup>2</sup>n (%) of total

**Figure 1.**
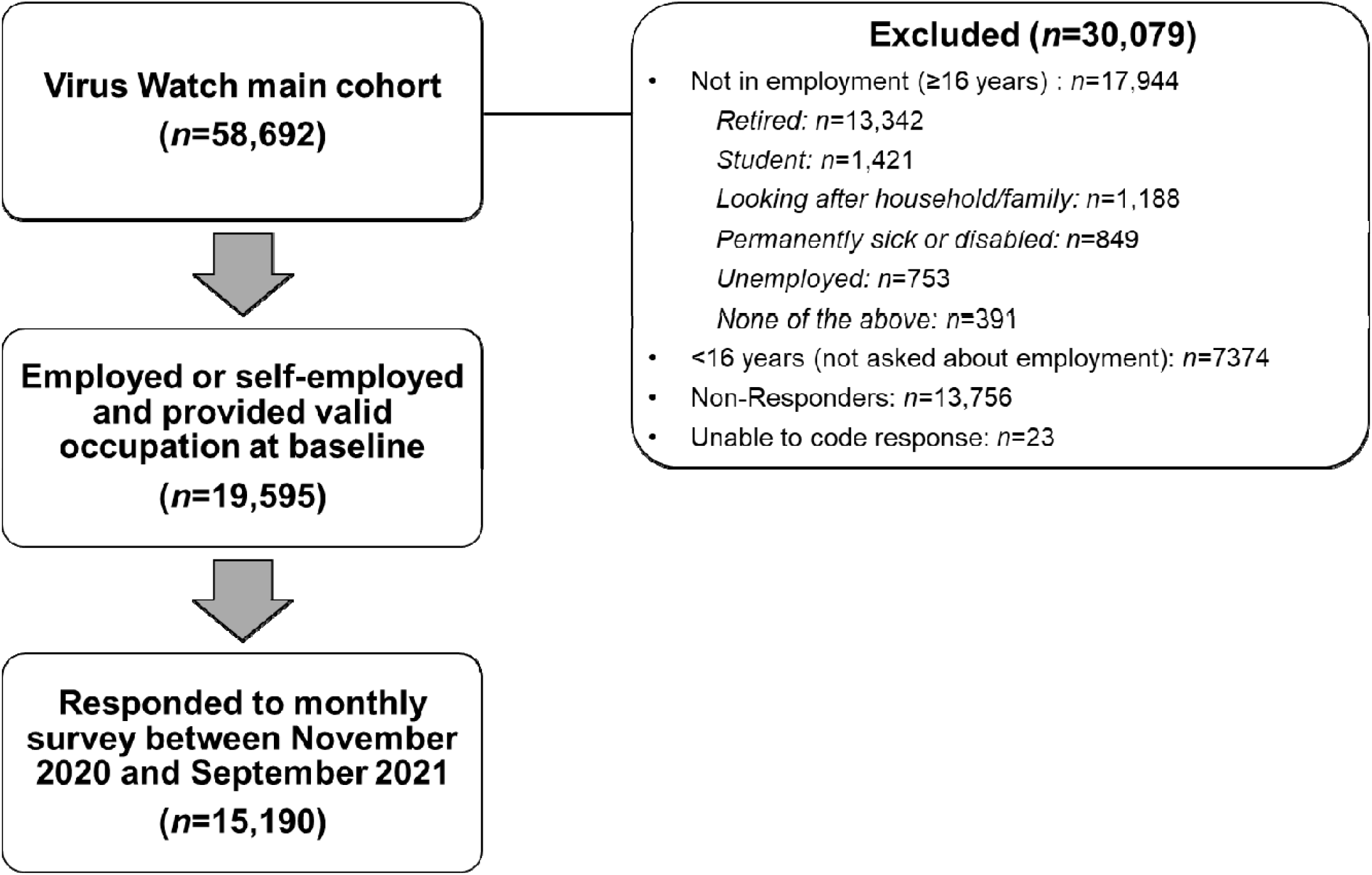
Flow Diagram of Participant Eligibility

### Occupational Group and Infection Risk

Absolute risk of infection by occupational risk is reported in Supplementary Table S2, and ranged from 26% in outdoor tradespeople to 42% in teaching, education and childcare workers across the full pandemic period covered by the study.

Across the full pandemic period covered, healthcare (adjusted risk ration (aRR)=1.29, 1.18-1.40; attributable fraction (AF) = 22%, 15-29%), leisure and personal service (aRR=1.15, 1.03-1.29; AF=, 13%, 3-22%), social care and community protective service (aRR=1.24, 1.12-1.38; AF=19%, 11-27%), and teaching, education and childcare occupations (aRR=1.34, 1.24-1.44; AF=25%, 19-30%) demonstrated elevated infection risk compared to Other Professional and Associate Occupations (Figure 2; see Supplementary Table S3 for adjusted AFs). When limited to participants who underwent serological testing (Supplementary Figure 2), similar groups demonstrated elevated infection risk with the addition indoor trade and process/plant workers (aRR=1.38, 1.07-1.78) and administrative and secretarial occupations (aRR=1.32, 1.06-1.64).

**Figure 2.**
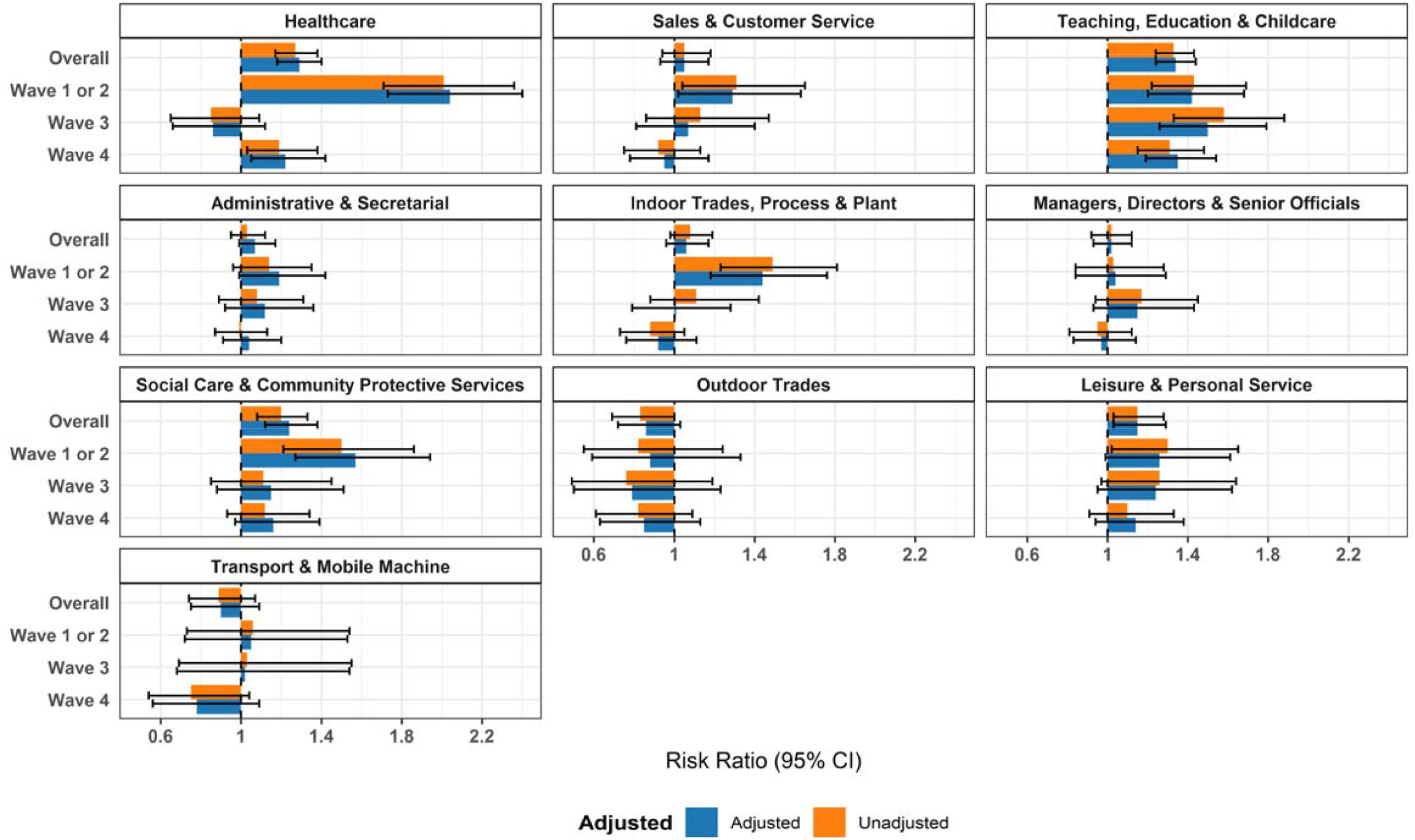
Risk Ratios by Occupational Group (versus Other Professional and Associate)

In Waves 1 and 2, healthcare (aRR=2.04, 1.73-2.40; AF=51%, 42-58%), indoors trades/process/plant (aRR=1.44, 1.18-1.76; AF=31%, 15-43%), sales and customer service (aRR=1.29, 1.02-1.94; AF=22%, 2-39%), social care and community protective services (aRR=1.57, 1.27-1.94; AF=36%, 21-48%), and teaching/education/childcare occupations (aRR=1.42, 1.20-1.68; AF=30%, 17-41%) demonstrated elevated risk. Only teaching, education and childcare occupations remained at elevated risk in Wave 3 (aRR=1.50, 1.26-1.79; AF=33%, 20-44%). Teaching, education and childcare workers were also at elevated risk in Wave 4 (aRR=1.35, 1.19-1.54; AF=26%, 16-35%), along with healthcare workers (aRR=1.22, 1.05-1.42; AF=18%, 5-30%). Across all models, limited effects of adjustment for sociodemographic, health-related and non-workplace activities were observed (Figure 2). Similar results were obtained in sensitivity analyses including imputed sociodemographic data (Supplementary Figure 3a).

Similar between-occupational trends were obtained when comparing each occupation to the rest of the working population (Figure 3 and Supplementary Table S2), with lower risk ratios and attributable fractions than those compared to Other Professional and Associate occupations. Similar results were also observed in related sensitivity analyses (Supplementary Figures 2 and 3b).

**Figure 3.**
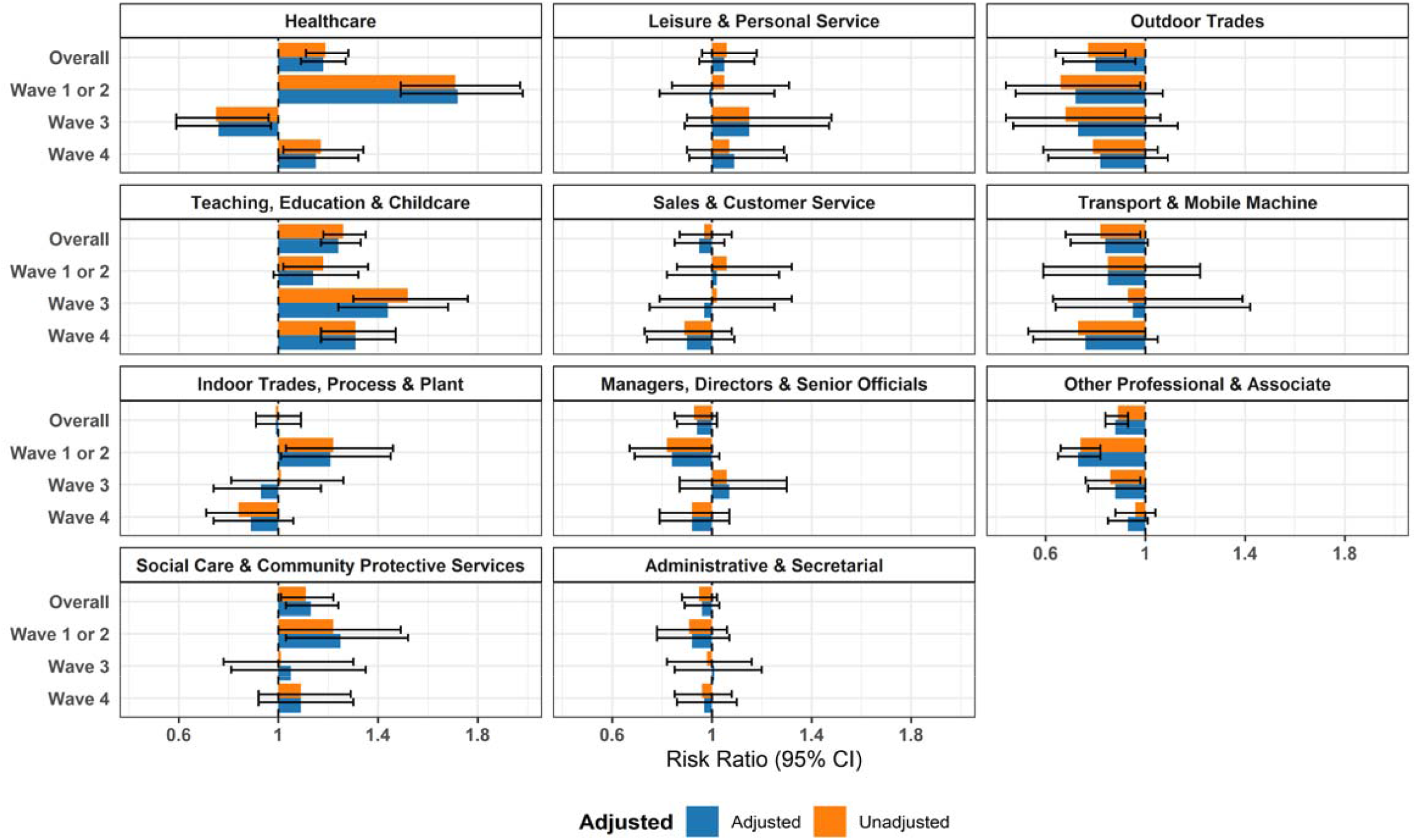
Risk Ratios by Occupational Group (versus Working Population)

### Specific Frontline Occupations

Absolute risk of infection for specific frontline occupations is reported in Supplementary Table S4; primary school teachers demonstrated the highest absolute risk (53%) across the full pandemic period.

The following frontline occupational groups demonstrated elevated infection risk compared to ‘Other professional and associate’ occupations: nurses (aRR=1.44, 1.25-1.65; AF=30%, 20-39%); doctors (aRR=1.33, 1.08-1.65; AF=25%, 7-39%); carers (1.45, 1.19-1.76; AF=31%, 16-43%); primary school teachers (aRR=1.67, 1.42-1.96; AF=40%, 30-49%); secondary school teachers (aRR=1.48, 1.26-1.72; AF=32%, 21-42%); and teaching support occupations (aRR=1.42, 1.23-1.64; AF=29%, 18-39%) (Figure 4; attributable fractions in Supplementary Table S5). All of these occupational groups demonstrated elevated risk during Waves 1 and 2, along with cleaners (aRR=1.60, 1.01-2.51; AF=37%, 1-60%); warehouse and process/plant workers (aRR=1.93, 1.41-2.65; AF=48%, 29-62%); and food preparation and hospitality workers (aRR=1.82, 1.17-2.83; AF=45%, 15-65%). In Wave 3, carers (aRR=1.91, 1.21-3.01; AF=48%, 18-67%), primary school teachers (aRR=1.72, 1.10-2.68; AF=42%, 9-63%), secondary school teachers (aRR=1.76, 1.21-2.56; AF=43%, 17-61%), and teaching support workers (aRR=1.70, 1.23-2.34; AF=41%,19-57%) demonstrated evidence of elevated risk. In Wave 4, primary school teachers (aRR=1.88, 1.39-2.53; AF=47%, 28-60%), secondary school teachers (aRR=1.52, 1.12-2.04; AF=34%, 11-51%), and teaching support workers (aRR=1.41, 1.09-1.83; AF=29%,8-45%) continued to demonstrate elevated risk. Similar results were obtained in sensitivity analyses with imputed sociodemographic data (Supplementary Figure 4a). Patterns of results were also similar when comparing frontline occupations to the rest of the working population (Figure 5 and Supplementary Table S4; sensitivity analyses in Supplementary Figure 4b), with attenuated risk ratios and attributable fractions than when using Other Professional and Associate occupations as a comparator.

**Figure 4.**
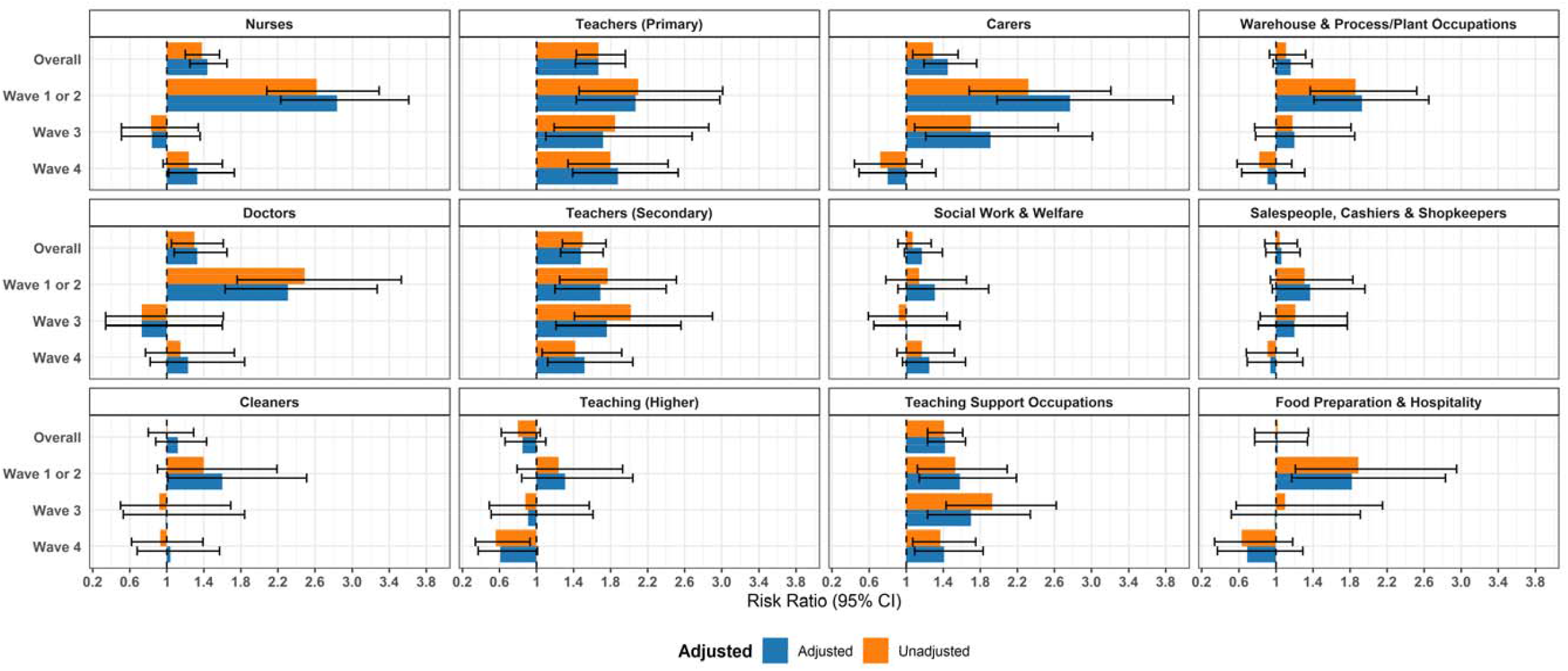
Risk Ratios for Frontline Occupations (versus Other Professional and Associate)

**Figure 5.**
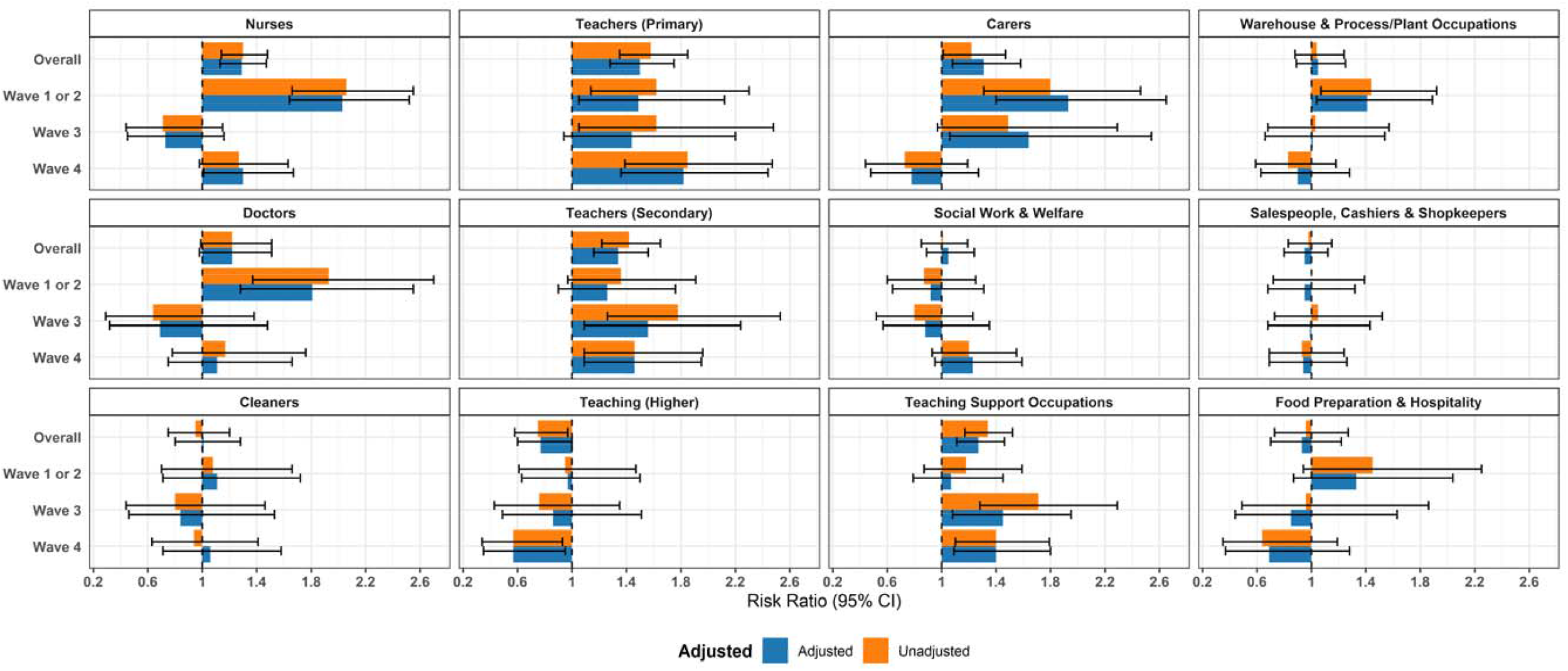
Risk Ratios for Frontline Occupations (versus Working Population)

## Discussion

### Key Findings and Interpretation

This study found persistent occupational differences in SARS-CoV-2 infection risk after comprehensive adjustment for non-work-related confounding, including socio-demographic and health-related factors and non-work social activities. Compared to Other Professional and Associate occupations - the largest occupational group in the sample with the lowest infection risk - workers in healthcare, teaching, education and childcare, social care and community protective services, and leisure and personal service occupations demonstrated elevated overall infection risk. In these groups, belonging to their occupation compared to the less risky group accounted for between 13% (for leisure and personal service workers) to 25% (for teaching, education and childcare workers) of their infection risk. Most of these at-risk occupations demonstrated elevated risk in the earlier pandemic phase (Waves 1 and 2) – along with indoor tradespeople and sales and customer service workers, who also demonstrated elevated risk during this period. This elevated relative risk was later attenuated for most occupational groups, with the exception of teaching, education and childcare occupations for whom risk remained elevated in Waves 3 and 4, and healthcare workers who also had elevated risk in Wave 4.

Where sample size was sufficient, we also investigated infection risk for specific frontline occupational groups. Nurses, doctors, carers, teachers, and teaching support workers demonstrated elevated risk compared to Other Professional and Associate occupations and – excluding the latter group – the rest of the working population across the full study period. Cleaners, warehouse and process/plant workers, and food preparation and hospitality workers also demonstrated evidence of elevated risk in Waves 1 and 2 only. Belonging to their occupation compared to Other Professional and Associated occupations accounted for between 25% (in doctors) to 40% (in primary school teachers) of at-risk workers’ risk of infection. Patterns of risk by pandemic phase were similar to those described above for broad occupational groups. Evidence of elevated risk for at-risk groups was most prominent in the early pandemic waves. Teachers and teaching support workers continued to demonstrate elevated risk in Waves 3 and 4, along with carers in Wave 3 and nurses in Wave 4. Findings may have been impacted by lack of power to detect modest effects in some groups.

Elevated infection risk in occupational groups with limited ability to work from home and those involving exposure to patients and/or the public echoes findings from the previous studies with more limited adjustment for potential confounding and from other global regions ^1 2 3 4 5 12 13^. Across all analyses in the current study, adjustment for sociodemographic and health-related factors and non-work activities had limited impact on estimates. This result differs markedly to prior analysis of occupational differences in COVID-19 mortality ^11^, where adjustment for socio-demographic and health-related factors substantially reduced the effect of occupation. Occupation plausibly shapes SARS-CoV-2 exposure - and consequently infection risk - by influencing workers’ ability to work from home, practise social distancing at work, work in well-ventilated environments, and access appropriate personal protective equipment. The specific mechanisms and relative contribution of different mitigating factors are likely to differ considerably by occupation, and are an important area for future research. Conversely, clinical factors that influence risk of severe morbidity and mortality once infected may differ across occupations, however the direct effect of occupation itself on severity of infection is likely to be more limited.

Changing patterns of differential infection risk by pandemic phase are likely to be multifactorial. Immunity-related factors that reduce the population of susceptible workers within a given occupation are likely to be important, and include prior infection in early phases of the pandemic, prioritization of some occupational groups (i.e. health and care workers ^11,26^) for vaccination, and potential differences in the speed and overall uptake of vaccination between occupations ^27^. The removal of remaining public health restrictions in Wave 3 may also have reduced differential risk by increasing overall contact rates and networks, and probability of transmission outside of work due to the increasing range of potential venues for exposure at a time of persistently high community infection rates and reduced mitigations. Resurgent risk in healthcare workers, particularly nurses, in the fourth wave may reflect the impact of relaxed restrictions on some healthcare workers with intensive patient contact as well as the impact of the immune-evasive Omicron variant. Relatedly, persistently elevated risk in teaching and childcare occupations may reflect high-intensity workplace exposure in combination with high levels of infection in children^28,29^. Direct investigation into potential mediators of this phase effect was beyond the scope of this study, and is warranted to better understand the processes shaping occupational infection risk. Relatedly, investigation into effective mitigation for the ongoing elevated infection risk in teachers is recommended both to address occupational inequalities and to reduce disruption in education settings.

### Strengths and Limitations

Strengths of this study include the large and diverse cohort that enabled investigation of infection risk from multiple study-derived and linked sources including both symptomatic testing and serology over multiple pandemic phases. Detailed information around participants’ demographic characteristics and activities over time allowed adjustment for a comprehensive series of potential confounders, including non-work-related public activities, informed by a directed acyclic graph.

However, the study has several important limitations. The Virus Watch cohort is demographically diverse but not representative of the UK population, with underrepresentation of some occupational categories limiting the ability to investigate differential risk across all occupational categories. Potential confounders, such as deprivation, are challenging to measure and residual confounding cannot be excluded. Non-work public activities were inferred from self-reported activities across a given survey week, and may not have been an accurate reflection of participants’ activity patterns across the entire relevant time period. Furthermore, social and leisure activities may have included work for some occupational groups (e.g. leisure and personal service occupations) but could not be disaggregated; however, the limited effect of adjustment in these models indicates that this was unlikely to be a major source of bias. Occupation was measured in broad categories, and only some specific occupations could be investigated due to small subsample sizes. Relatedly, the number of infections within a given pandemic phase was small for some frontline subsamples. Overall estimates of risk by occupational sector may be driven by particularly risky roles with considerable exposure ^9^, and further investigation into specific occupations is recommended. Additionally, inclusion of multiple test types to indicate SARS-CoV-2 positivity allowed for potential detection of asymptomatic or previously untested cases through serology, and detection of early cases through linkage. However, issues impacting the uptake and usage of each test type, including differential access to some tests in given phases of the pandemic, self-selection bias, and compliance with testing instructions may have affected estimates and are difficult to delineate. Notably, swab testing uptake may be influenced by differential testing behaviour between occupations. For example, health care workers undertake regular occupational testing which may lead to an overestimation of their relative risk of infection. However, a sensitivity analysis constrained to those participants who underwent serological testing was not subject to such testing behaviour bias and demonstrated similar results to the main analyses.

## Conclusions

Despite these limitations, the present study indicates differential infection risk across occupational groups in England and Wales, with patterns of differential risk appearing to vary across pandemic phase. These findings illustrate the importance of work as a source of infection risk during the COVID-19 pandemic, with substantial fractions of infections attributable to occupation in at-risk groups. Occupations with persistently elevated risk (i.e. teachers) should be an ongoing target for interventions such as improved ventilation in schools, while understanding processes that shape differential risk in earlier phases of the pandemic is relevant for future outbreaks of respiratory infections. Investigation into the mechanisms underlying differential risk overall and over time, as suggested by this study, could inform evidence-based public health interventions in the workplace.

## Data Availability

We aim to share aggregate data from this project on our website and via a "Findings so far" section on our website - https://ucl-virus-watch.net/. We also share some individual record level data on the Office of National Statistics Secure Research Service. In sharing the data we will work within the principles set out in the UKRI Guidance on best practice in the management of research data. Access to use of the data whilst research is being conducted will be managed by the Chief Investigators (ACH and RWA) in accordance with the principles set out in the UKRI guidance on best practice in the management of research data. We will put analysis code on publicly available repositories to enable their reuse.

