## Supplementary Material for "Differential Risk of SARS-CoV-2 Infection by Occupation: Evidence from the Virus Watch prospective cohort study in England and Wales"

**Supplementary Materials**

**Supplementary Table 1. UK Standard Occupational Classification 2020 (SOC-2020) Codes within Virus Watch Occupational Categories**

| **Virus Watch Occupational Category** | **UK SOC-2020 Codes** | **Three Most Prevalent Occupations***  **(SOC-2020 Unit Group)** |
| --- | --- | --- |
| Administrative & Secretarial Occupations | 4111-4217, 9211, 9219, 9233 | 1. Other administrative occupations n.e.c. (21%, n= 328) 2. Book-keepers, payroll managers, and wage clerks (11%, n=168) 3. National government administrative occupations (7%, n=111) |
| Healthcare Occupations | 2211-2259, 3211-3219, 3240, 6131-6133 | 1. Other nursing professionals (21%, n=193) 2. Generalist medical practitioners (11%, n=101) 3. Nursing auxiliaries and assistants (6%, n=55) |
| Indoor Trades, Process & Plant Occupations | 5211-5250, 5315-5317, 5321-5323, 5411-5449, 8111-8149, 8160, 9131-9139, 9241-9259 | 1. Warehouse operatives (10%, n=85) 2. Metalworking production and maintenance fitters (8%, n=65) 3. Chefs (7%, n=56) |
| Leisure & Personal Service Occupations | 1221-1225, 1252, 1253, 1256, 1257, 6121, 6129, 6211-6250, 9221-9229, 9231, 9261-9269 | 1. Cleaners and domestics (14%, n=82) 2. Kitchen and catering assistants (8%, n=45) 3. Security guards and related occupations (6.4%, n=37) |
| Managers, Directors & Senior Officials | 1111-1161, 1171,1172, 1211, 1212, 1231, 1241-1243, 1251, 1254, 1255, 1258, 1259 | 1. Financial managers and directors (16%, n=161) 2. Human resource managers and directors (9.9%, n=99) 3. Functional managers and directors n.e.c. (9.6%, n=96) |
| Other Professionals & Associate Professionals | 2111-2162, 2411-2455, 2471-2494, 3111-3133, 3411-3582 | 1. Programmers and software development professionals (5.5%, n=222) 2. Business and financial project management professionals (4%, n=161) 3. Management consultants and business analysts (3.8%, n=152) |
| Outdoor Trade Occupations | 5111-5119, 5311-5314, 5319, 5330, 8151-8159, 9111- 9129 | 1. Gardeners and landscape gardeners (24%, n=61) 2. Construction and building trades n.e.c. (20%, n=50) 3. Construction operatives n.e.c. (12%, n=30) |
| Sales & Customer Service Occupations | 7111-7220 | 1. Sales and retail assistants (36%, n=227) 2. Customer service occupations n.e.c. (12%, n=74) 3. Sales supervisors - retail and wholesale (11%, n=68) |
| Social Care & Community Protective Services | 1162, 1163, 1232, 2461-2469, 3221-3229, 3311-3319, 6134-6138, 6311-6312 | 1. Care workers and home carers (26%, n=160) 2. Welfare and housing associate professionals n.e.c. (12%, n=75) 3. Police officers (sergeant and below) (9%, n=55) |
| Teaching, Education & Childcare Occupations | 1233, 2311-2329, 3231, 3232, 6111-6117, 9232 | 1. Secondary education teaching professionals (14%, n=196) 2. Education advisers and school inspectors (14%, n=185) 3. Higher education teaching professionals (13%, n=177) |
| Transport & Mobile Machine Operatives | 8211-8239 | 1. Large goods vehicle drivers (21%, n=56) 2. Delivery drivers and couriers (20%, n=54) 3. Bus and coach drivers (14%, n=37) |

**Abbreviations:** n.e.c. = not elsewhere classified; * Limited to three most prevalent occupations per category to prevent declarative disclosure and due to large number of occupations across sample (*n*=402)

**Clinical Outcomes**

**Ascertainment**

Infection during the study period was defined as participants’ first positive result from any of the following sources of reporting:

1) Polymerase chain reaction (PCR) or lateral flow (LFT) tests either self-reported by the whole Virus Watch cohort, taken as part of the study (see study protocol for details^1^) or obtained from linkage of all participants’ data to the UK national laboratory reporting system
2) Serological positives: Virus Watch monthly at-home finger-prick serology

or in-clinic serology. In-clinic serology was conducted twice per participants between September 2020-January 2021 (Autumn round) and April 2021-July 2021 (Spring round) (see study protocol for details^1^). 9114 included participants received at-home and/or in-clinic serology.

Sliding date window matching was used to match any positive tests recorded by both Virus Watch and linkage to UK national records within the same 14-day window; in these cases, the linked date was used. Where both PCR or LFT positives and serological positives were available, we used the PCR/LFT date to assign a wave, unless the serological positive occurred first. As this analysis pertained to non-household transmission, we restricted cases where possible to those acquired outside of the household (e.g., no other individual in the household with a positive PCR or lateral flow test or, where there were multiple cases in the household, the first case in the household based on earliest date of symptom onset). This was not possible for participants detected through serological testing.

**Result Source**

Of the 5236 positive results, 4275 (82%) were ascertained based on swab tests and 961 (18%) on serology.

**Supplementary Figures 1a and 1b. Directed Acyclic Graphs for Occupation – Infection Risk**

**1a**

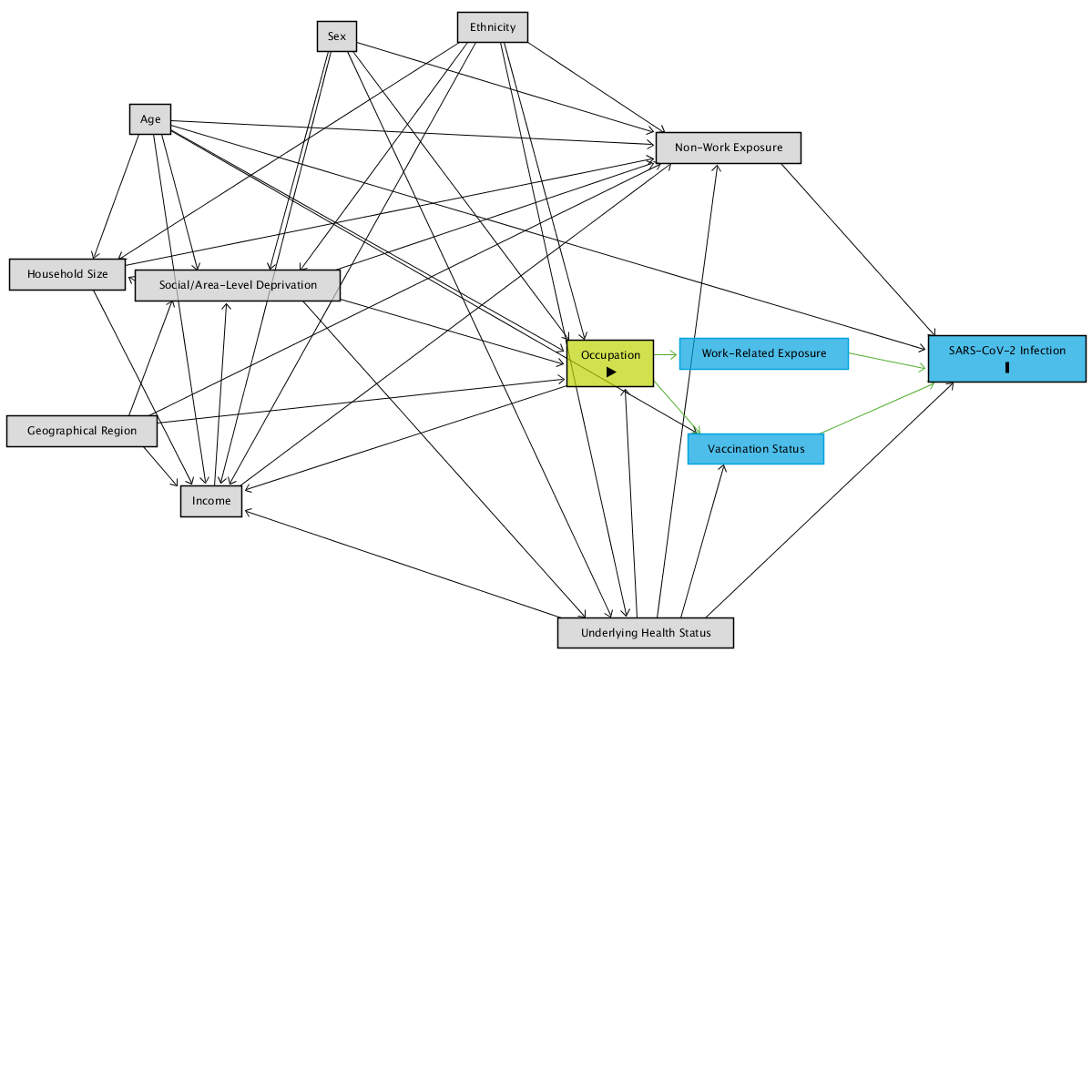

**1b**

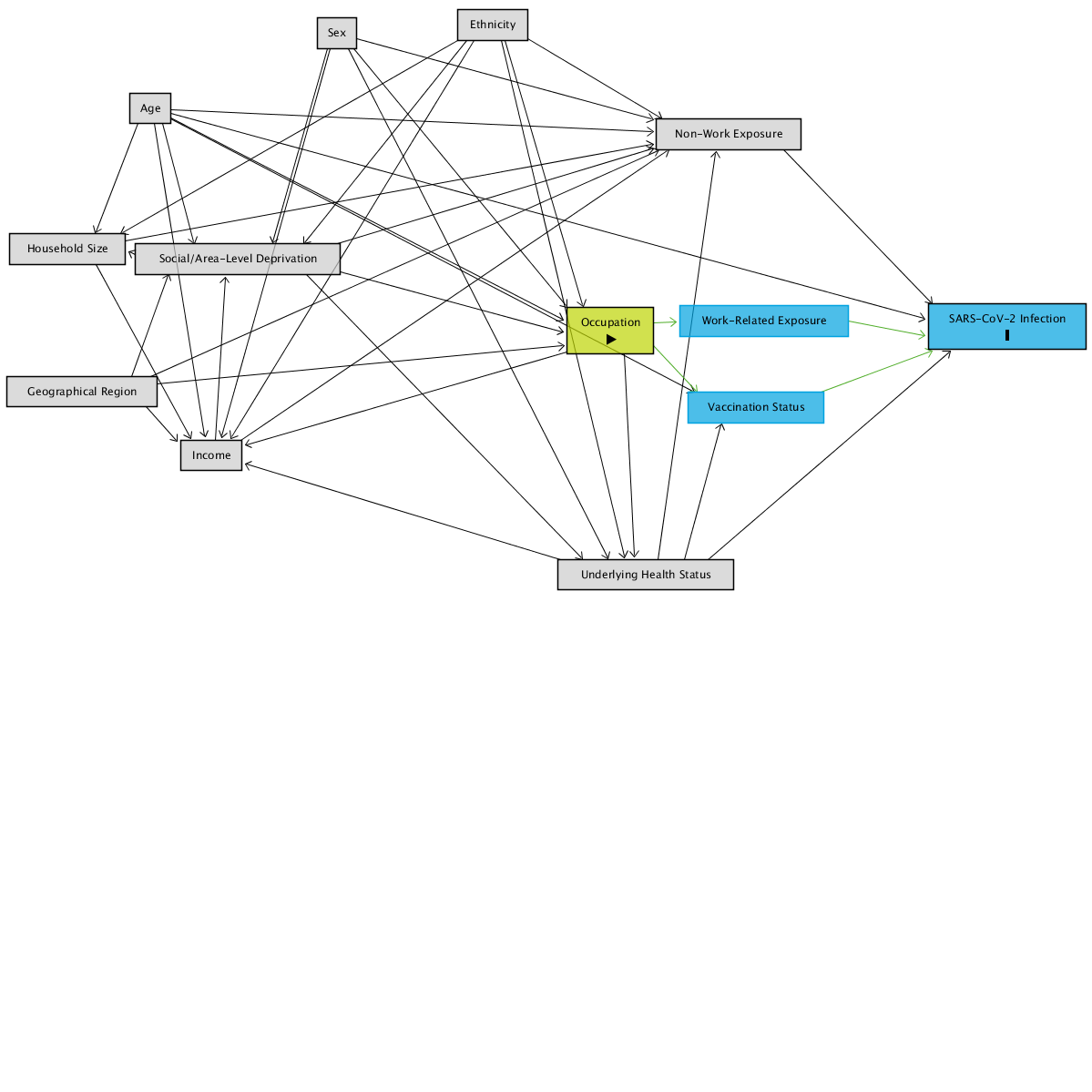

**Note:** Supplementary Figure 1a and 1b illustrate how the current adjustment set remains sufficient under assumptions of health status influencing occupation or occupation influencing health status. Black arrows denote paths controlled by adjustment, while green arrows denote causal paths.

**Supplementary Table 2. Absolute Infection Risk by Occupational Group**

|  |  | N | Total Positive* |  |  | Wave 1 or 2 Positive* |  | Wave 3 Positive* | Wave 4 Positive* |
| --- | --- | --- | --- | --- | --- | --- | --- | --- | --- |
| Administrative & Secretarial |  | 1,942 | 650 (33%) |  |  | 184 (9.5%) |  | 161 (8%) | 285 (15%) |
| Healthcare |  | 1,272 | 518 (41%) |  |  | 204 (16%) |  | 72 (6%) | 211 (17%) |
| Indoor Trades, Process & Plant |  | 1,044 | 357 (34%) |  |  | 124 (12%) |  | 83 (8%) | 133 (13%) |
| Leisure & Personal Service |  | 731 | 266 (36%) |  |  | 85 (12%) |  | 64 (9%) | 109 (15%) |
| Managers, Directors & Senior Officials |  | 1,249 | 409 (33%) |  |  | 102 (8%) |  | 107 (9%) | 186 (15%) |
| Other professional & associate |  | 4,972 | 1,575 (32%) |  |  | 422 (8.5%) |  | 362 (7%) | 739 (15%) |
| Outdoor Trades |  | 372 | 96 (26%) |  |  | 26 (7%) |  | 19 (5%) | 46 (12%) |
| Sales & Customer Service |  | 770 | 253 (33%) |  |  | 84 (11%) |  | 61 (8%) | 102 (13%) |
| Social Care & Community Protective Services |  | 827 | 315 (38%) |  |  | 98 (12%) |  | 65 (8%) | 131 (16%) |
| Teaching, Education & Childcare |  | 1,671 | 702 (42%) |  |  | 204 (12%) |  | 186 (11%) | 290 (17%) |
| Transport & Mobile Machine |  | 340 | 96 (28%) |  |  | 29 (8.5%) |  | 23 (7%) | 39 (11%) |
| **Total** |  | 15,190 | 5,237 (34%) |  |  | 1,562 (10%) |  | 1,203 (8%) | 2,271 (15%) |

*Proportion of occupational category total

**Supplementary Table 3. Attributable Fractions (Exposed) by Occupational Group**

|  | **versus Other Professional and Associate – AF (95% CI)** | | | | **versus Working Population – AF (95% CI)** | | | |
| --- | --- | --- | --- | --- | --- | --- | --- | --- |
|  | **Overall** | **Wave 1 or 2** | **Wave 3** | **Wave 4** | **Overall** | **Wave 1 or 2** | **Wave 3** | **Wave 4** |
| Administrative & Secretarial | 6.86 (-1.14,14.22) | 15.81 (-0.54,29.50) | 10.34 (-8.97,26.23) | 4.27 (-9.81,16.55) | -4.15 (-12.04,3.18) | -9.08 (-27.43,6.63) | 0.80 (-17.87,16.51) | -3.24 (-16.78,8.73) |
| Healthcare | 22.27 (15.42,28.57) | 50.96 (42.17,58.42) | -16.13 (-51.17,10.78) | 18.24 (4.90,29.71) | 15.02 (8.34,21.21) | 41.71 (32.81,49.42) | -32.16 (-69.07,-3.31) | 13.19 (0.31,24.40) |
| Indoor Trades, Process & Plant | 5.90 (-3.99,14.85) | 30.61 (15.35,43.11) | 0.50 (-26.92,22) | -8.86 (-31.05,9.57) | -0.62 (-10.44,8.32) | 17.58 (1.30,31.18) | -7.50 (-35.08,14.44) | -12.82 (-34.59,5.42) |
| Leisure & Personal Service | 13.00 (2.64,22.26) | 20.74 (-1.10,37.86) | 19.50 (-4.97,38.27) | 12.19 (-6.45,27.57) | 4.84 (-5.72,14.35) | -0.58 (-26.17,19.81) | 12.73 (-11.82,31.89) | 8.01 (-10.29,23.28) |
| Managers, Directors & Senior Officials | 2.16 (-7.61,11.04) | 4.19 (-18.73,22.69) | 12.98 (-8.02,29.90) | -3.01 (-20.85,12.2) | -6.84 (-16.79,2.26) | -19.14 (-45.48,2.44) | 6.32 (-14.53,23.37) | -9.17 (-26.94,6.10) |
| Other Professional & Associate | **Ref** | **Ref** | **Ref** | **Ref** | -13.63 (-19.76,-7.81) | -36.2 (-52.68,-21.50) | -13.71 (-29.13,-0.12) | -8.1 (-18.00,0.96) |
| Outdoor Trades | -16.25 (-39.77,3.31) | -13.38 (-70.59,24.64) | -27.3 (-100.04,18.99) | -18.34 (-58.51,11.65) | -25.17 (-49.95,-4.48) | -39.83 (-108.58,6.26) | -37.64 (-114.21,11.56) | -22.33 (-62.85,8.10) |
| Sales & Customer Service | 4.34 (-7.28,14.69) | 22.52 (1.92,38.80) | 6.35 (-23.07,28.73) | -4.80 (-28.53,14.55) | -5.57 (-17.56,5.19) | 1.90 (-21.99,21.1) | -3.10 (-33.05,20.12) | -11.69 (-35.56,7.97) |
| Social Care & Community Protective Services | 19.42 (10.59,27.38) | 36.24 (21.06,48.50) | 13.19 (-13.55,33.63) | 13.77 (-3.51,28.16) | 11.54 (2.60,19.66) | 19.93 (2.61,34.18) | 4.51 (-22.8,25.75) | 8.54 (-8.58,22.96) |
| Teaching, Education & Childcare | 25.17 (19.40,30.53) | 29.76 (16.90,40.62) | 33.35 (20.44,44.16) | 26.02 (15.89,34.94) | 19.52 (14.16,24.54) | 12.08 (-1.75,24.04) | 30.59 (19.12,40.43) | 23.49 (14.21,31.76) |
| Transport & Mobile Machine | -11.01 (-33.96,8.01) | 4.47 (-39.52,34.59) | 1.93 (-47.91,34.98) | -27.92 (-77.63,7.88) | -19.21 (-43.26,0.80) | -18.08 (-70.68,18.31) | -4.84 (-56.45,29.75) | -31.65 (-81.70,4.61) |

**Supplementary Figure 2. Risk Ratios by Occupational Group – Serologically-Confirmed Infections vs No Evidence of Infection**

a. ***Risk Ratios versus Other Professional and Associate Occupations***
b. ***Risk Ratios versus Working Population***

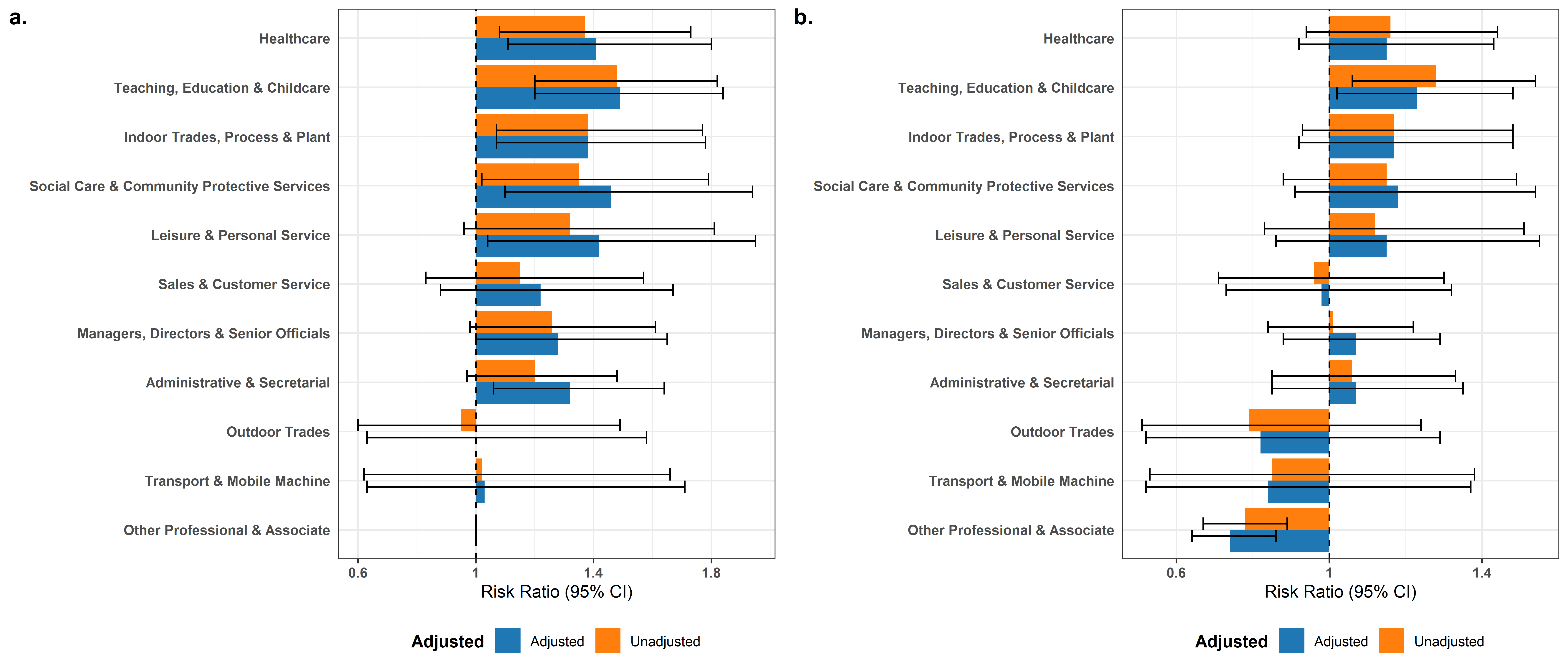

**Supplementary Figure 3a and 3b. Risk Ratios by Occupational Group – Missing Data Sensitivity Analyses with Imputation**

a. ***Risk Ratios versus Other Professional and Associate Occupations***

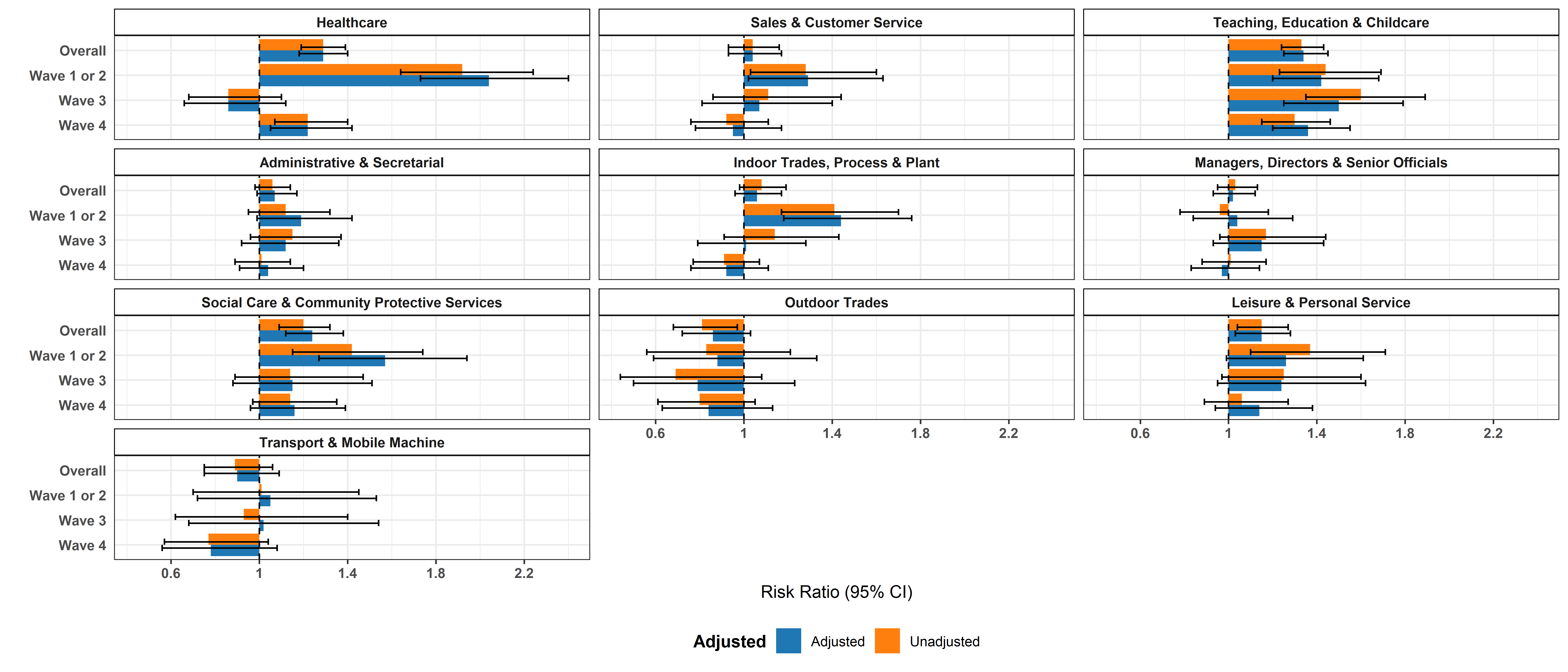

b. ***Risk Ratios versus Working Population***

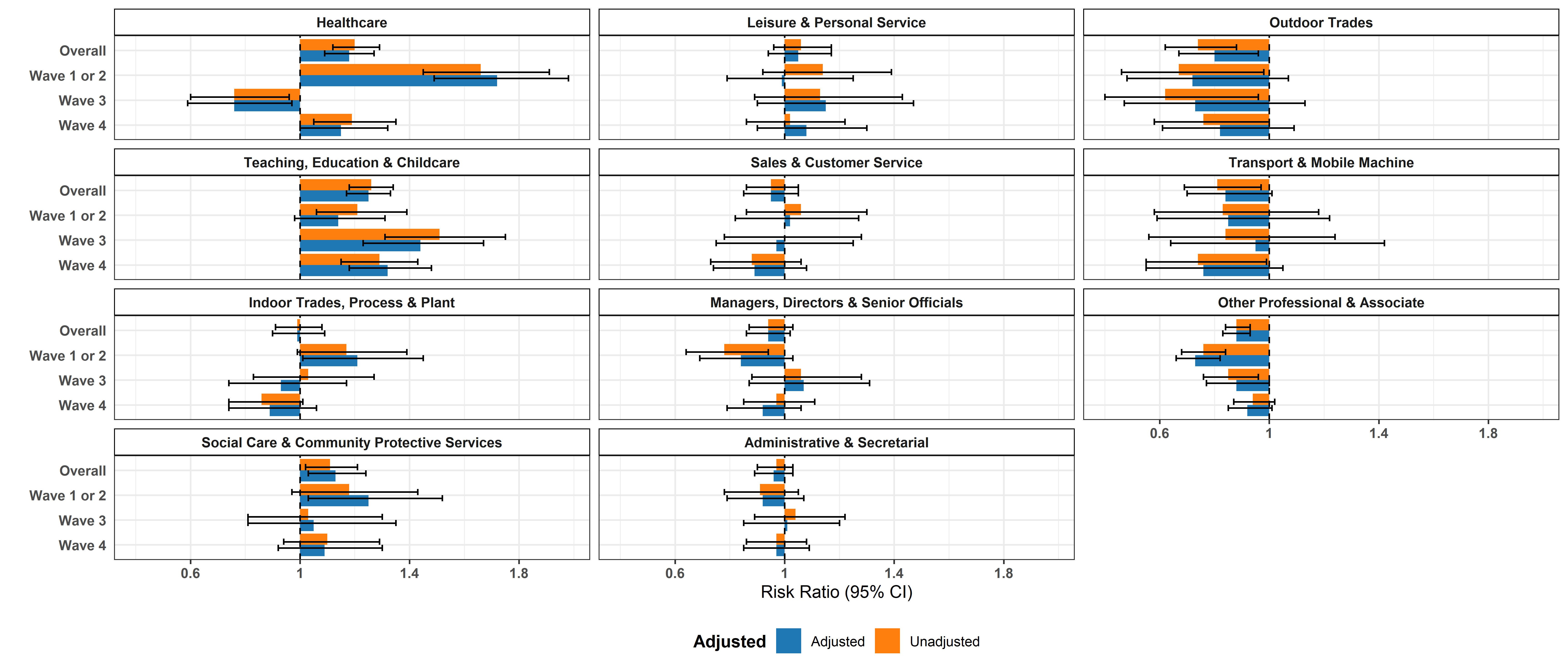

**Supplementary Table 4. Absolute Infection Risk for Frontline Occupations**

| **Characteristic** | **N** | **Total Positive*** |  | **Wave 1 or 2 Positive*** |  | **Wave 3 Positive*** | **Wave 4 Positive*** |
| --- | --- | --- | --- | --- | --- | --- | --- |
| Carers | 162 | 67 (41%) |  | 32 (20%) |  | 18 (11%) | 14 (9%) |
| Cleaners | 148 | 48 (32%) |  | 18 (12%) |  | 10 (7%) | 20 (13.5%) |
| Doctors | 125 | 52 (42%) |  | 27 (22%) |  | 6 (5%) | 19 (15%) |
| Food Preparation & Hospitality | 104 | 34 (33%) |  | 17 (16%) |  | 8 (8%) | 9 (9%) |
| Nurses | 300 | 132 (44%) |  | 68 (23%) |  | 16 (5%) | 48 (16%) |
| Salespeople, Cashiers, & Shopkeepers | 291 | 97 (33%) |  | 33 (11%) |  | 26 (9%) | 38 (13%) |
| Social Work & Welfare | 274 | 94 (34%) |  | 27 (10%) |  | 19 (7%) | 48 (17.5%) |
| Teachers (Higher) | 168 | 43 (26%) |  | 18 (11%) |  | 11 (6.5%) | 14 (8%) |
| Teachers (Primary) | 144 | 77 (53%) |  | 26 (18%) |  | 18 (12.5%) | 32 (22%) |
| Teachers (Secondary) | 190 | 91 (48%) |  | 29 (15%) |  | 27 (14%) | 34 (18%) |
| Teaching Support Occupations | 286 | 129 (45%) |  | 38 (13%) |  | 40 (14%) | 51 (18%) |
| Warehouse & Process/Plant Occupations | 242 | 86 (36%) |  | 39 (16%) |  | 20 (8%) | 27 (11%) |

*Proportion of occupational category total

**Supplementary Table 5. Attributable Fractions (Exposed) for Frontline Occupations**

|  | **versus Other Professional and Associate – AF (95% CI)** | | | | **versus Working Population – AF (95% CI)** | | | |
| --- | --- | --- | --- | --- | --- | --- | --- | --- |
|  | **Overall** | **Wave 1 or 2** | **Wave 3** | **Wave 4** | **Overall** | **Wave 1 or 2** | **Wave 3** | **Wave 4** |
| Nurses | 30.45 (20.25,39.34) | 64.76 (55.17,72.29) | -19.30 (-94.22,26.72) | 24.86 (2.13,42.32) | 22.50 (11.87,31.86) | 50.78 (38.85,60.39) | -37.82 (-121.25,14.15) | 23.03 (1.02,40.15) |
| Doctors | 24.91 (7.01,39.37) | 56.63 (38.51,69.41) | -36.16 (-195.86,37.34) | 18.71 (-21.35,45.54) | 17.98 (-1.55,33.76) | 44.66 (22.02,60.73) | -45.25 (-212.50,32.49) | 10.30 (-33.83,39.87) |
| Cleaners | 10.68 (-13.76,29.87) | 37.39 (1.46,60.22) | -1.23 (-88.54,45.64) | 3.54 (-46.29,36.40) | 1.00 (-25.01,21.60) | 9.83 (-39.90,41.89) | -19.35 (-117.83,34.61) | 5.43 (-41.72,36.89) |
| Salespeople, Cashiers, & Shopkeepers | 5.30 (-12.57,20.33) | 27.20 (-3.90,48.99) | 16.71 (-22.88,43.54) | -6.04 (-44.71,22.30) | -5.39 (-24.41,10.71) | -5.82 (-47.93,24.31) | -1.28 (-46.86,30.16) | -6.75 (-43.94,20.83) |
| Carers | 30.95 (16.24,43.08) | 63.96 (49.59,74.23) | 47.69 (17.60,66.79) | -24.37 (-104.23,24.26) | 23.51 (7.78,36.56) | 48.14 (28.82,62.21) | 39.15 (5.82,60.68) | -28.41 (-109.50,21.30) |
| Social Work & Welfare | 14.38 (-1.65,27.88) | 23.55 (-10.43,47.07) | 1.47 (-53.61,36.80) | 20.14 (-4.51,38.97) | 4.91 (-12.20,19.41) | -9.05 (-55.55,23.56) | -13.95 (-74.72,25.68) | 18.62 (-5.40,37.17) |
| Teachers (Primary) | 40.10 (29.58,49.04) | 51.68 (30.30,66.50) | 41.77 (9.06,62.71) | 46.73 (28.14,60.51) | 33.15 (21.93,42.75) | 32.96 (4.70,52.84) | 30.36 (-6.89,54.62) | 45.03 (26.24,59.04) |
| Teachers (Secondary) | 32.28 (20.94,42.00) | 40.94 (16.32,58.31) | 43.16 (17.30,60.93) | 34.02 (11.02,51.07) | 25.49 (13.47,35.84) | 20.57 (-11.23,43.27) | 35.99 (8.42,55.27) | 31.59 (8.53,48.84) |
| Teachers (Higher) | -17.63 (-52.04,9.00) | 23.64 (-18.72,50.89) | -10.02 (-94.67,37.82) | -63.95 (-170.62,0.67) | -29.06 (-66.41,-0.09) | -2.86 (-58.85,33.39) | -15.67 (-102.64,33.98) | -73.99 (-186.17,-5.79) |
| Teaching Support Occupations | 29.52 (18.53,39.03) | 36.83 (12.62,54.34) | 41.14 (19.03,57.22) | 29.21 (8.13,45.46) | 21.50 (10.06,31.48) | 6.42 (-27.02,31.06) | 31.27 (7.79,48.76) | 28.81 (8.60,44.54) |
| Warehouse & Process/Plant Occupations | 13.97 (-2.83,28.03) | 48.23 (29.11,62.20) | 16.66 (-28.64,46.00) | -9.86 (-57.76,23.50) | 5.18 (-12.64,20.19) | 28.84 (4.07,47.22) | 1.15 (-50.46,35.06) | -11.34 (-59.07,22.07) |
| Food Preparation & Hospitality | 1.71 (-29.60,25.45) | 45.12 (14.85,64.63) | -0.52 (-92.95,47.64) | -44.68 (-170.90,22.73) | -7.97 (-41.89,17.85) | 25.06 (-14.59,51.00) | -17.78 (-125.63,38.52) | -45.65 (-172.44,22.13) |

**Supplementary Figure 4a and 4b. Risk Ratios for Frontline Occupations – Missing Data Sensitivity Analyses with Imputation**

***a. Risk Ratios versus Other Professional and Associate Occupations***

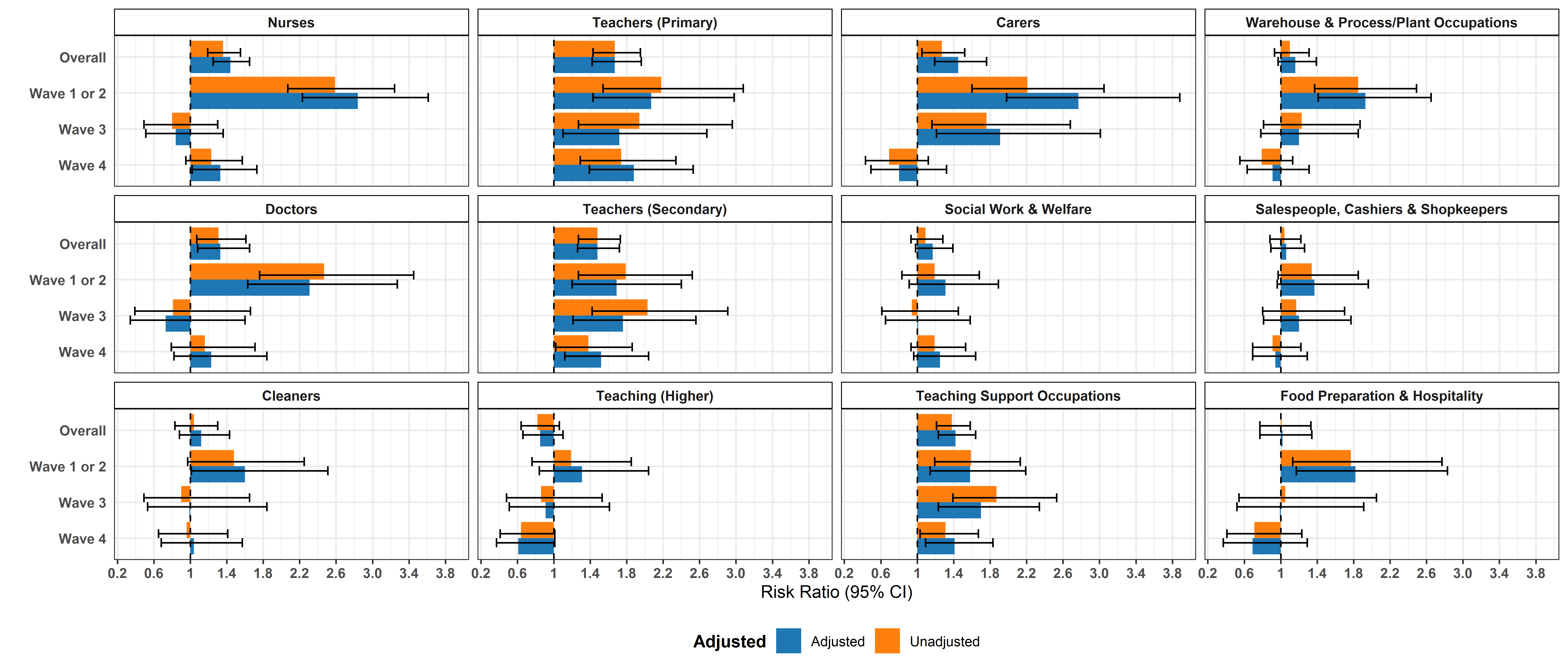

***b. Risk Ratios versus Working Population***

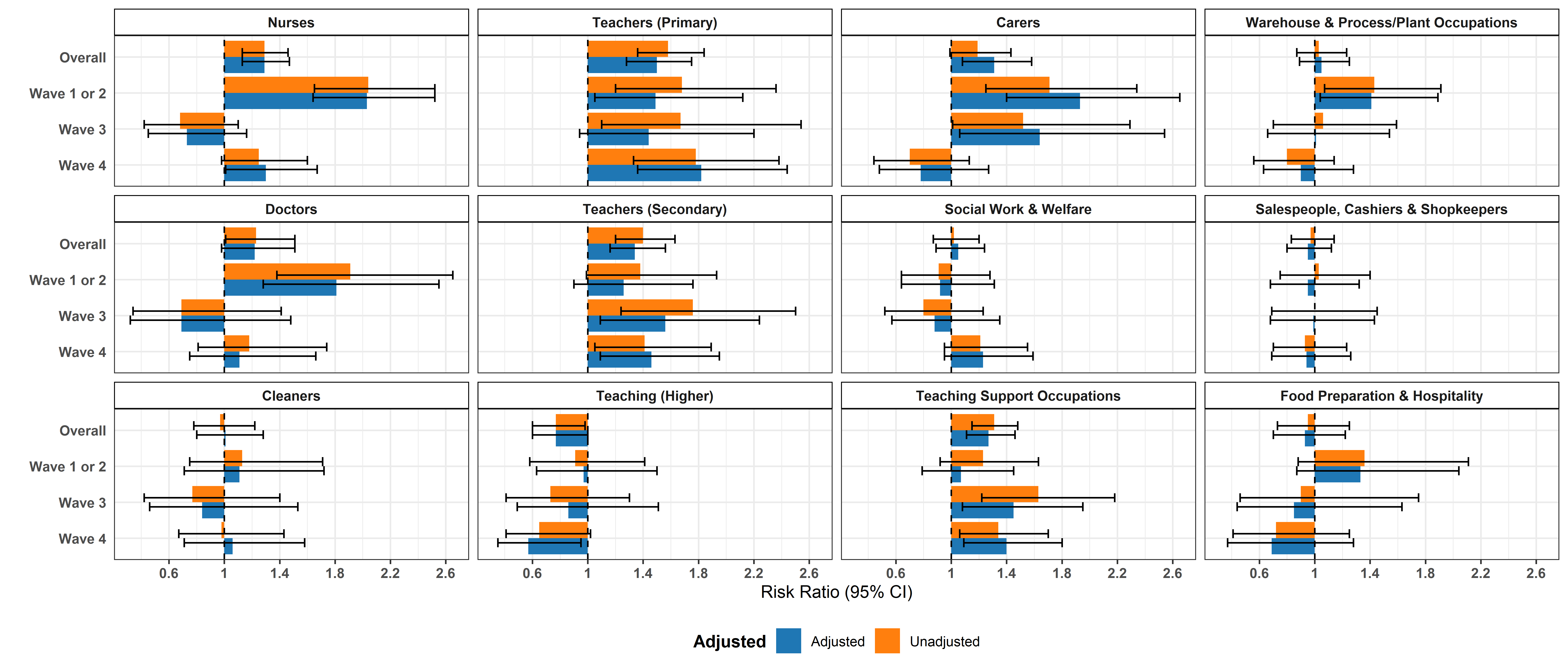
